# Examining COVID19 Positivity-Ratio Trends in US States from April-July: Are Rising Caseloads Attributable to ‘More Testing’ and Do State Political-Affiliations Play a Role?

**DOI:** 10.1101/2020.07.20.20158485

**Authors:** Bisakha P. Sen, Sangeetha Padalabalanarayanan

**Affiliations:** Department of Health Services Administration, University of Alabama at Birmingham, Birmingham, AL, USA; School of Health Professions, University of Alabama at Birmingham, Birmingham, AL, USA; Department of Health Care Organization and Policy, University of Alabama at Birmingham, Birmingham, AL, USA

**Author notes:** **Corresponding Author** Bisakha P. Sen, PhD, University of Alabama at Birmingham, RPHB 330, 1665 University Blvd, Birmingham, AL 35294-0022.

## Abstract

**Importance:** Rising Covid19 cases in the US are attributed by some political leaders to more testing. Positivity-ratios (cases to tests ratio) in conjunction with cases and tests provide a better overview. However, comprehensive overviews of positivity-ratio patterns are scarce.

**Objective:** To examine trends in positivity-ratios, tests and cases by state from mid-April-mid-July. Further, to examine whether positivity-ratio patters are associated with state political-affiliations.

**Methods:** State-level publicly available data on Covid19 is used. Seven-day moving averages (MA7) of positivity-ratio are computed for April 21-July 15. States are assigned to four groups based on patterns of change in positivity-ratio – higher at end of study period than beginning (Group 1), initial decline but subsequent increase starting Memorial Day weekend (Group 2), initial decrease but an upturn in last 14 days (Group 3), and consistent downward trend (Group 4). ‘Political-affiliation’ is categorized as ‘Republican-leaning’ if President Trump won the state and the governor is Republican. Additionally, a proxy measure is used to indicate intensity of Black Lives Matter (BLM) protests in the state. Associations are tested using chi-square analysis.

**Results:** Fourteen states are in Group 1, fifteen states in Group 2, fifteen states in Group 3, and six states and DC in Group 4. 78.57% of Group 1, 33.33% of Group 2, 40% of group 3, and none in Group 4 were Republican-leaning. The difference in distribution was statistically significant (p<0.01). Distribution of ‘high’ intensity BLM protests across the four groups was not statistically different (p>0.10).

**Conclusion:** Increased Covid19 cases cannot be attributed to more testing. Indeed, the high positivity-ratios in most states indicate current testing is failing to capture actual infection rates. The association between state political-affiliation and positivity-ratios suggests Republican voters may be somewhat more skeptical of the gravity of the disease and emphasizes the importance of messaging by political leaders.

## Introduction

Several U.S. states have seen sharp increases in COVID19 cases in the past several weeks. President Trump and Vice-President Pence have repeatedly argued that the growing number of (detected) cases is largely attributable to expanded testing^1-3^, and are balking at providing funds for additional testing^4^. However, public-health experts caution that the numbers reflect a surge in new infections, possibly due to inadequate adherence to safety measures like social-distancing and masks. After initial stay-at-home orders were lifted, there was anecdotal evidence of large-scale ignoring of safety measures during the Memorial Day weekend, and that pattern has pushed some states to consider re-instating partial shutdowns again.

One way to infer whether increases in reported COVID19 cases are attributable to more testing is to examine ‘positivity-ratios’ --the ratio of positive-cases detected to tests conducted. If underlying infection-rates are not rising, then when testing is expanded to include those deemed low-risk or presenting few symptoms, the positivity-ratio should decline even though case counts per se increase. A low positivity-ratio is one indicator that disease-spread is under control. In May, the World Health Organization recommended a positivity-ratio of 5% or lower for 2 weeks as a benchmark for re-opening economies^5,6^. However, while daily case-counts and trends are routinely reported in large media outlets and state health department websites, comprehensive information on trends in positivity-ratios are less readily accessible.

We provide a state-by-state and US as a whole overview in trends in positivity-ratio from April 21^st^-July 15^th^, thus covering a period that starts before states started relaxing stay-at-home orders, up to recent days. In conjunction, we provide trends in cases and tests. We believe this comprehensive picture helps dissipate some of the confusion around the extent to which more cases are attributable to more testing. We believe this will help inform public-health messaging, as well as the debate about the best way to approach school re-openings in the fall in different states.

We also explore whether patterns of changes in positivity-ratios correlate to a state’s political-affiliation. There is evidence of a ‘politicization’ of attitudes around the disease, with studies showing that those who identify as Republicans, or primarily follow conservative news outlets, are more likely to believe that risks of the virus are exaggerated ^7,8^. However, case counts are increasing sharply not just in states like Texas and Arizona, but also in Democratic states like California and Oregon. To our knowledge, no study has yet explored whether there are associations between positivity-ratios and state political affiliation. Finally, we are aware of concerns that the extensive Black Lives Matter (BLM) protests following Mr. George Floyd’s death^9^ – which occurred immediately after the Memorial Day weekend --may have also exacerbated virus spread. Hence, we use a proxy variable to capture how intense the protests were in each state, and explore the association between that and positivity-ratio patterns.

## Methods

Data on Covid19 positive-cases and tests are obtained from ‘The COVID Tracking Project’ (TCTP)^10^, which collates data from state health agencies and makes it publicly available. We construct positivity-ratios for April 21-July 15 using the ratio of 7-day-moving-average (MA7) of daily cases to MA7 of daily tests. MA7 is computed using data from the prior 6 and current date. If there are ‘negative counts’ of daily testing – which represent a correction for prior over counting, then we include them in our calculations with no manipulations. Positivity-ratio graphs are presented for each state and for the U.S., fractional polynomial regressions are used to estimate fitted trends, and corresponding graphs for daily MA7 for cases and tests are presented. Briefly, if there are time-periods when cases and tests are increasing but positivity-ratios are either declining or staying constant, then the argument can be made that more testing is finding more cases, but the underlying infection-rates are not climbing. However, if positivity-ratios are climbing, then this strongly indicates that underlying infection-rates are climbing, regardless of more tests.

States are categorized into four groups based on Positivity-ratio patterns. States with a higher positivity-ratios for July 13-15 than April 21-23 are categorized as Group 1. Of the rest, states with an initial decline but a subsequent increase so that positivity-ratios for July 13-15 are higher than Memorial Day weekend are categorized as Group 2. Of the rest, states with a largely downward trend but a marked upturn in the final 2-weeks are Group 3. The remaining states with a consistently downward trend over the study-period are Group 4. The categories are mutually exclusive by construction.

Political-affiliation is operationalized as ‘Republican leaning’ if President Trump won the state in the 2016 election and the current state-governor is Republican, and ‘not Republican leaning’ otherwise. Note that the ‘not Republican leaning’ includes states that have a Democratic governor and President Trump did not win the state, as well as states which meet either one of those conditions.

We use a proxy-measure for BLM protest-intensity based on actions taken by states as of June 4, when the protests were growing in momentum. Specifically, if curfews were instated, the National Guard was mobilized, and a state of emergency declared as well --then protest-intensity is defined as ‘high’. If all three did not happen, then it is ‘not high’^11^.

The relationship between positivity-ratio groups, political-affiliation and protest-intensity is tested using Chi-square analysis, and sensitivity of results examined by conducting Fisher’s Exact test. To examine sensitivity, we repeated the Chi-square analyses after excluding DC, Georgia, Hawaii, South Carolina and Washington since they have repeated negative values of MA7 for testing.

## Results

Figure 1 shows trends in positivity-ratios, cases and tests for the US overall and each state separately. According to this Figure, testing has increased monotonically for the US as a whole, but there was only a brief period in early June when cases were increasing while positivity-ratios were largely constant, and since mid-June positivity-ratios for the country as a whole have been increasing monotonically. The state-level graphs also indicate that positivity-ratios ae increasing for most states. There are only occasional examples --such as in Colorado, Delaware and Michigan – where it can be credibly argued that for most of the study period more cases are being detected because of expanded testing, since we see cases and tests increase with no concurrent increase in positivity-ratios.

**Figure 1.**
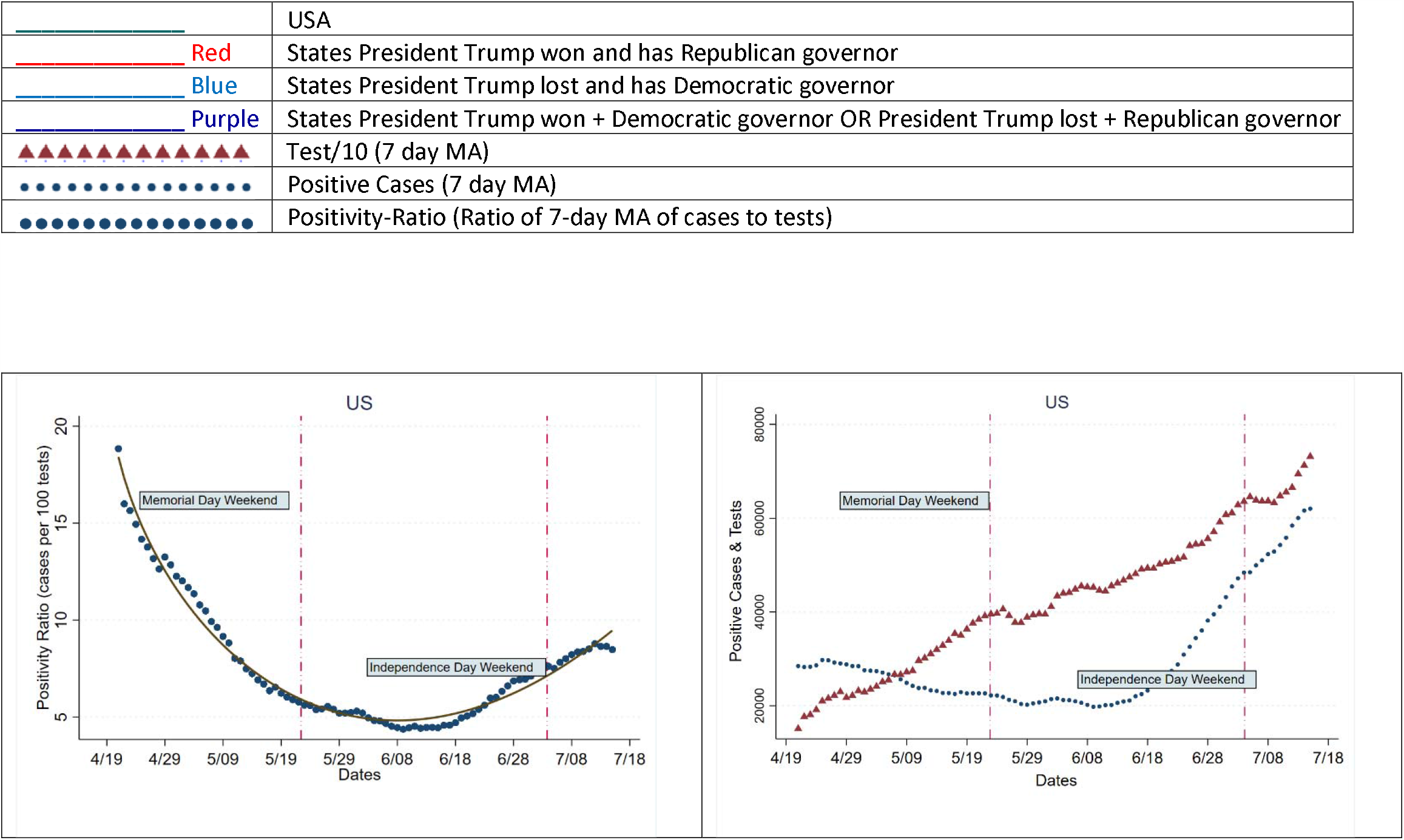
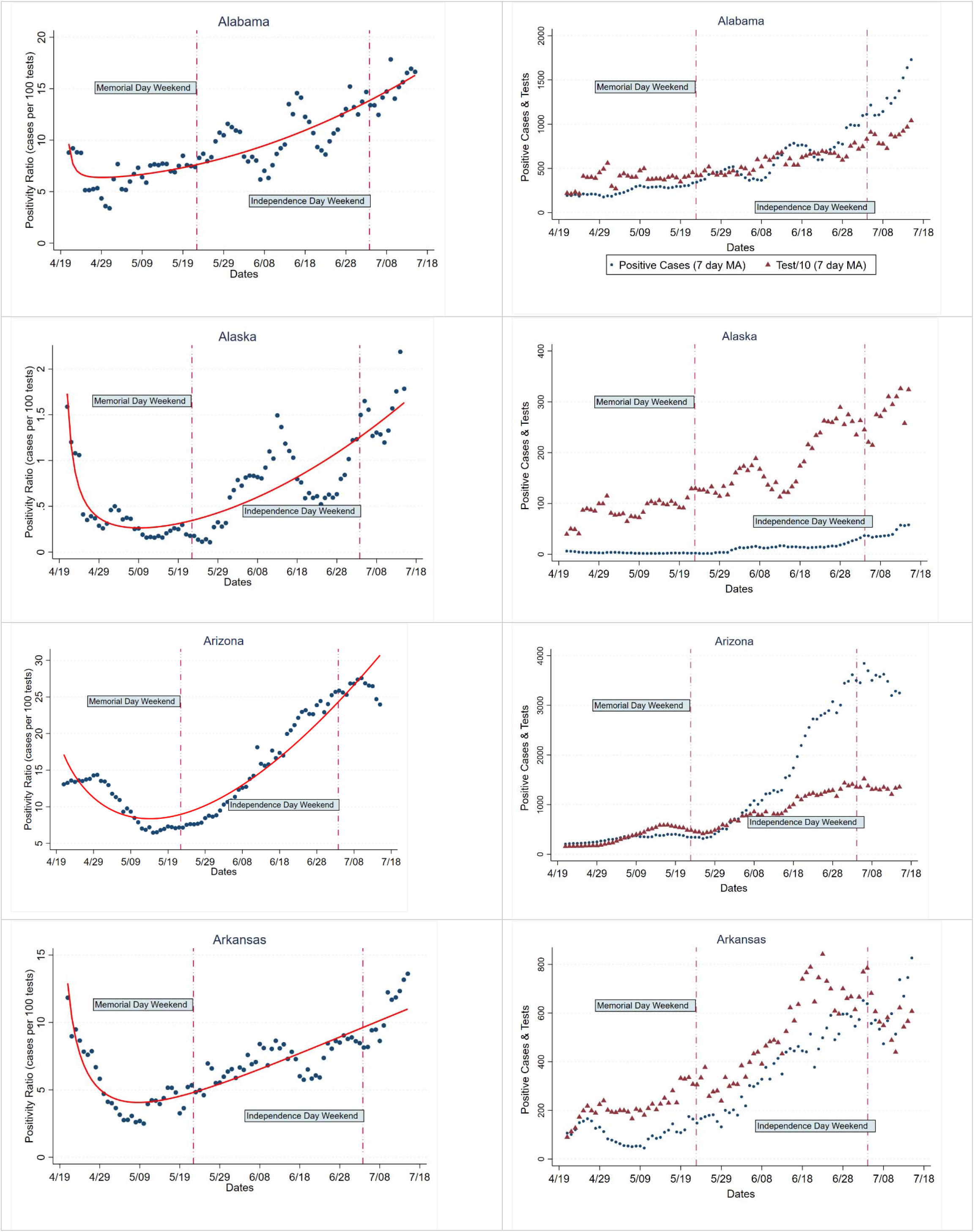
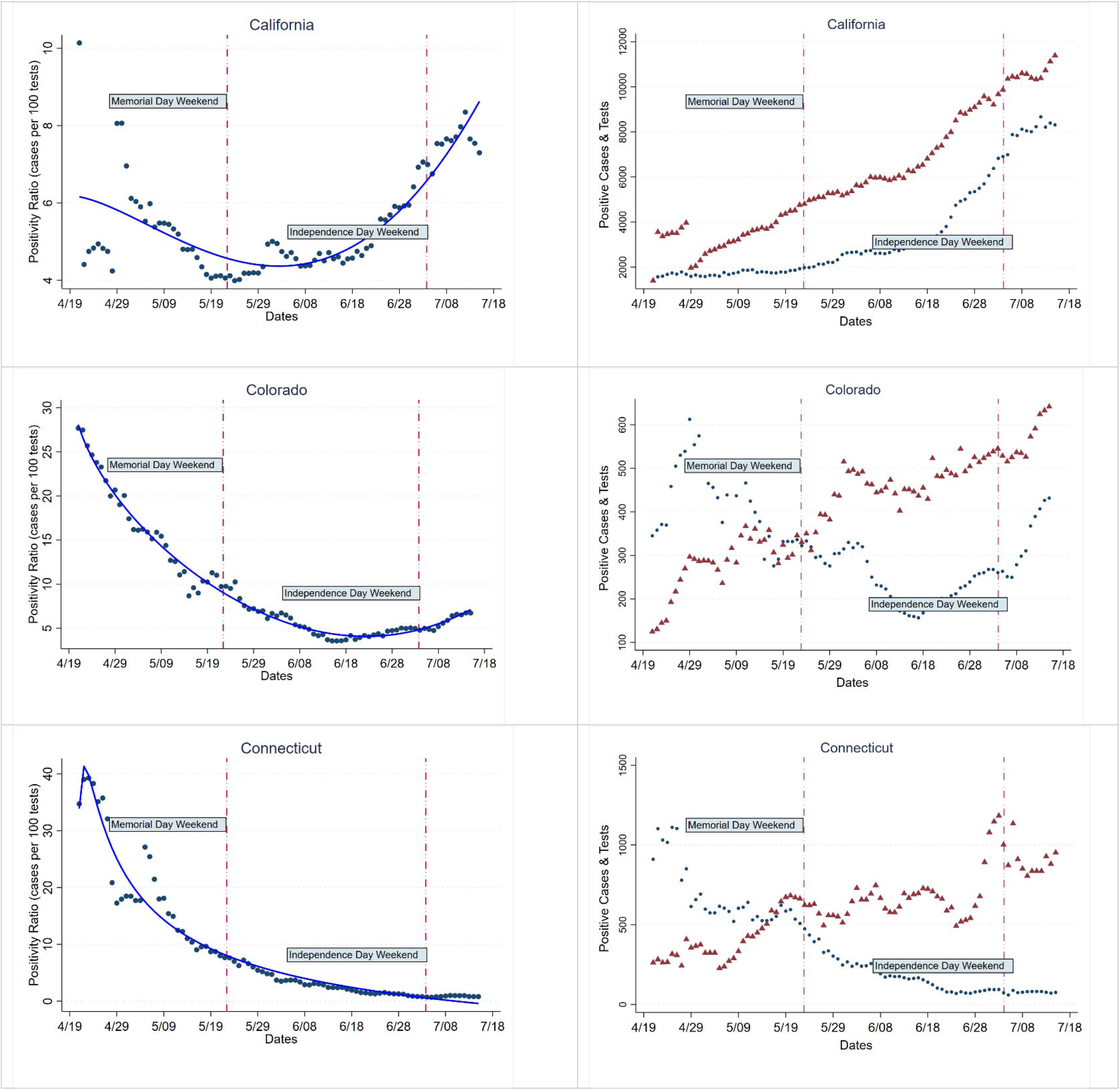
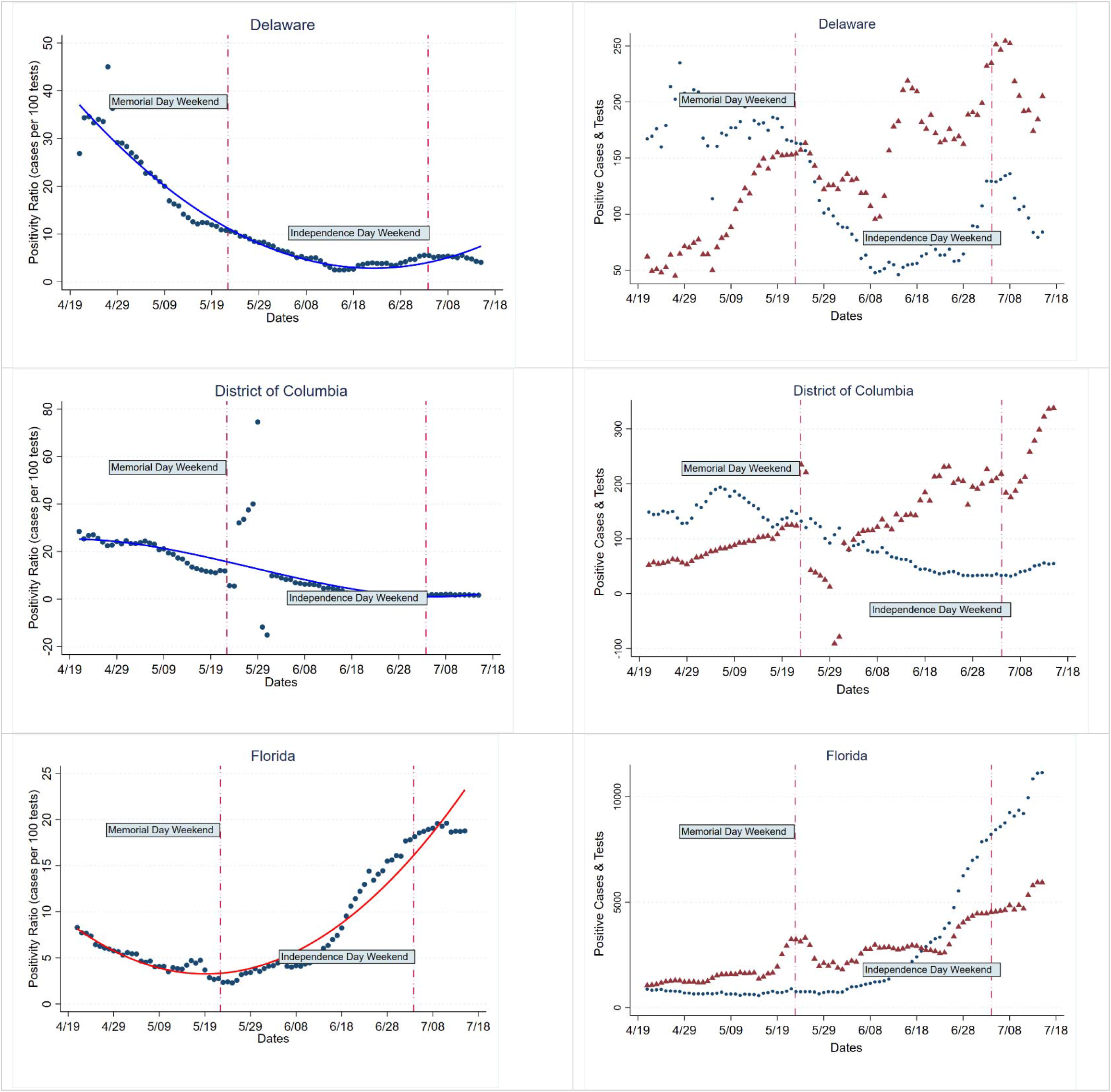
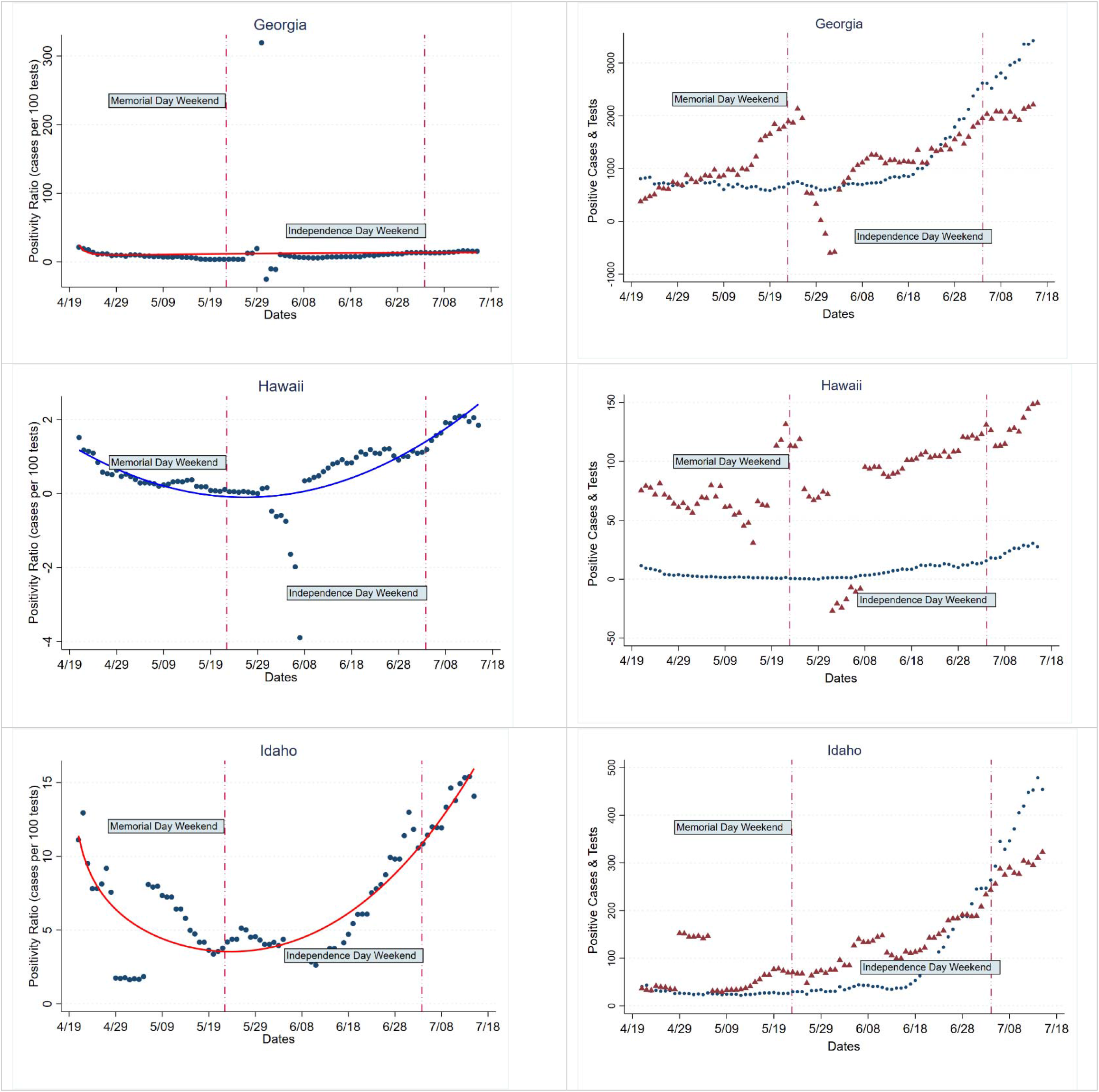
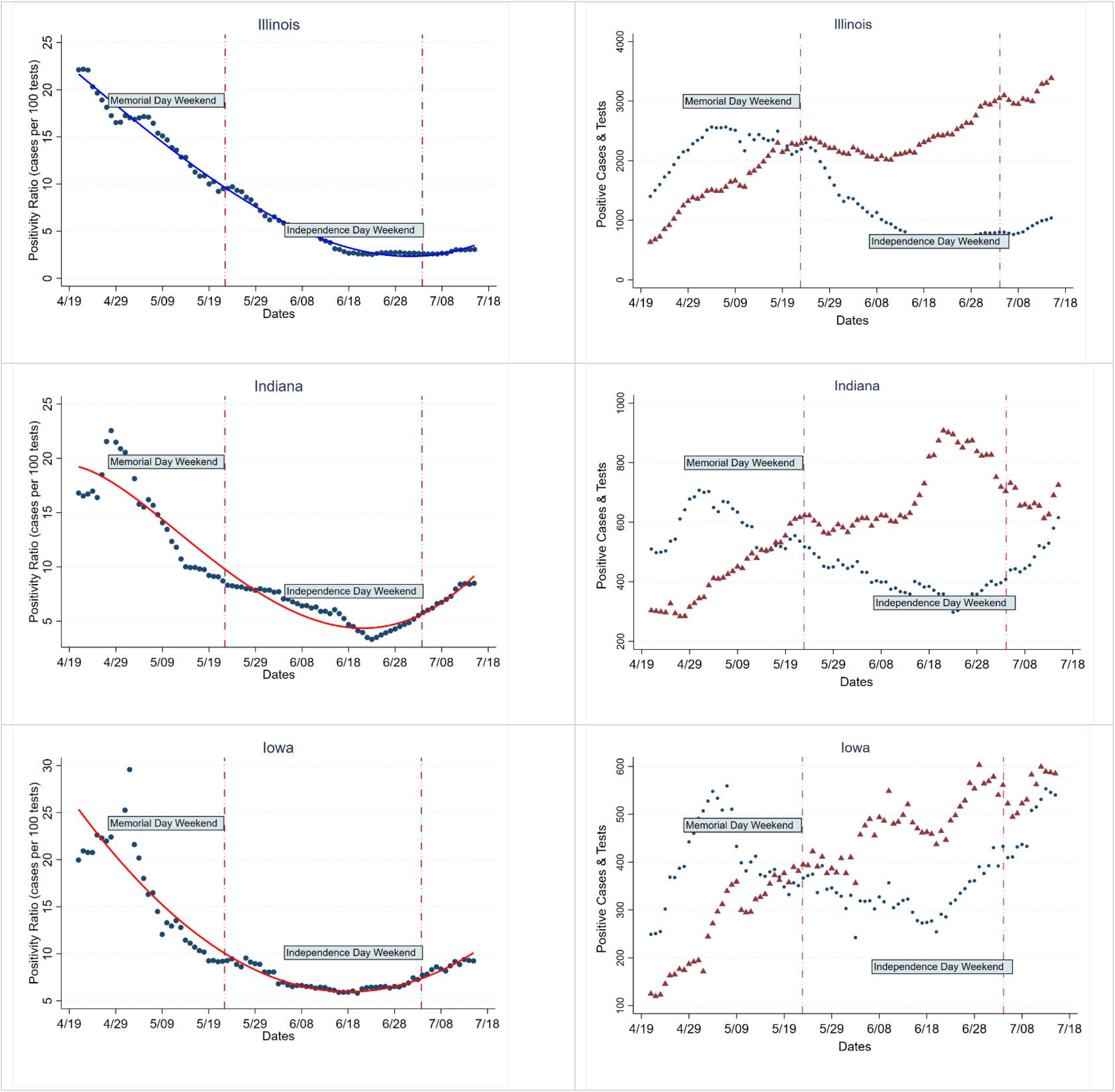
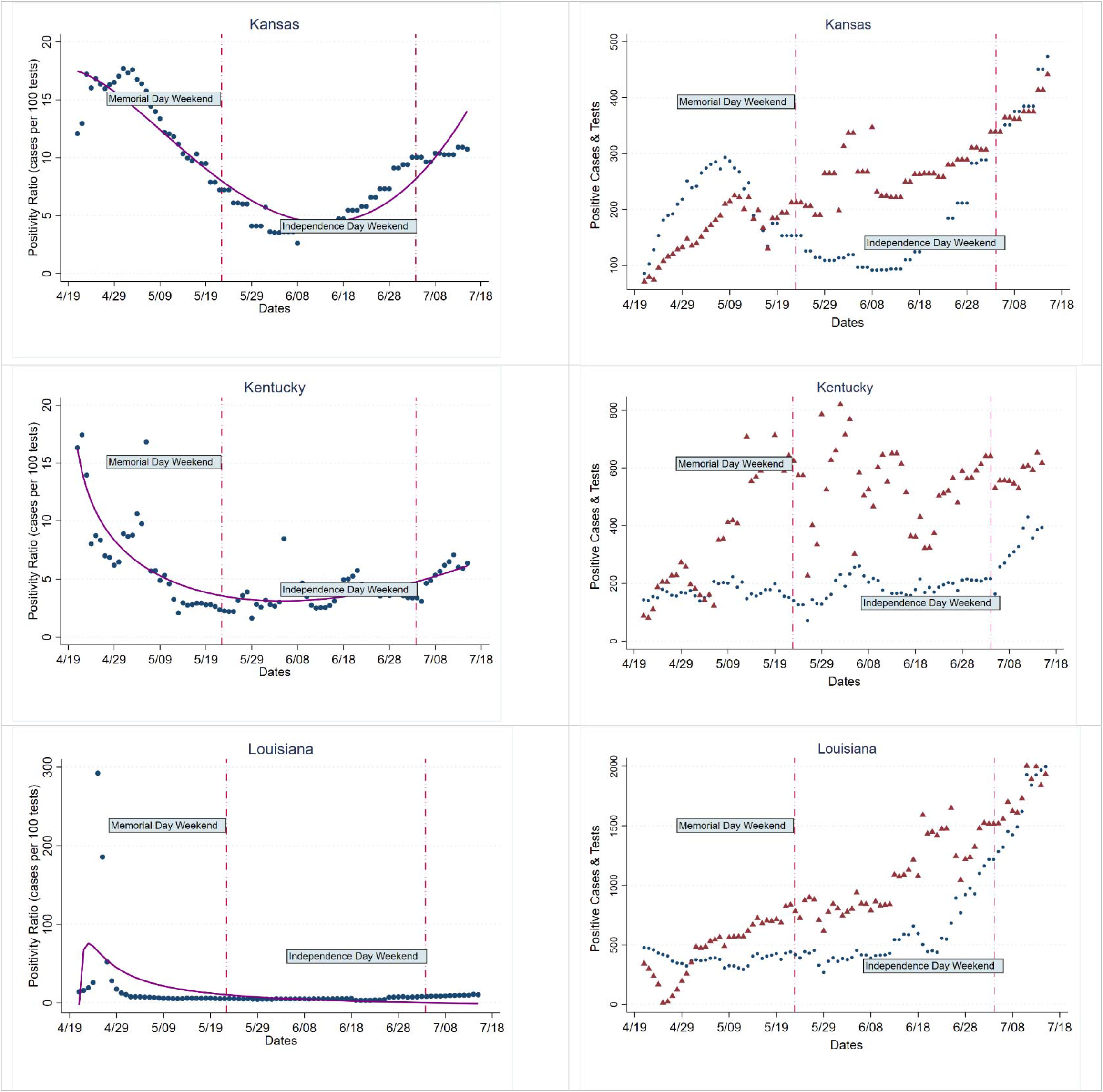
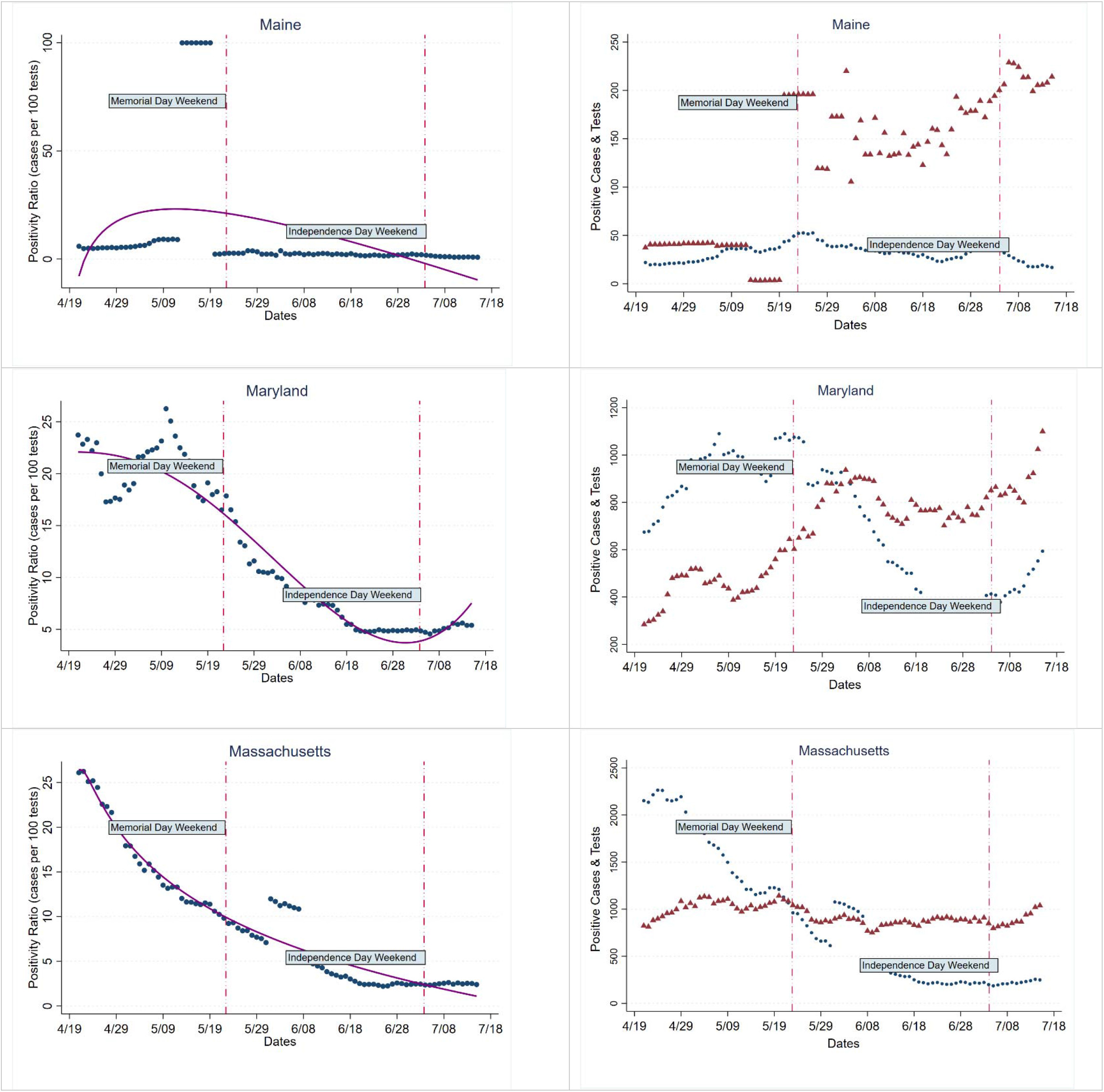
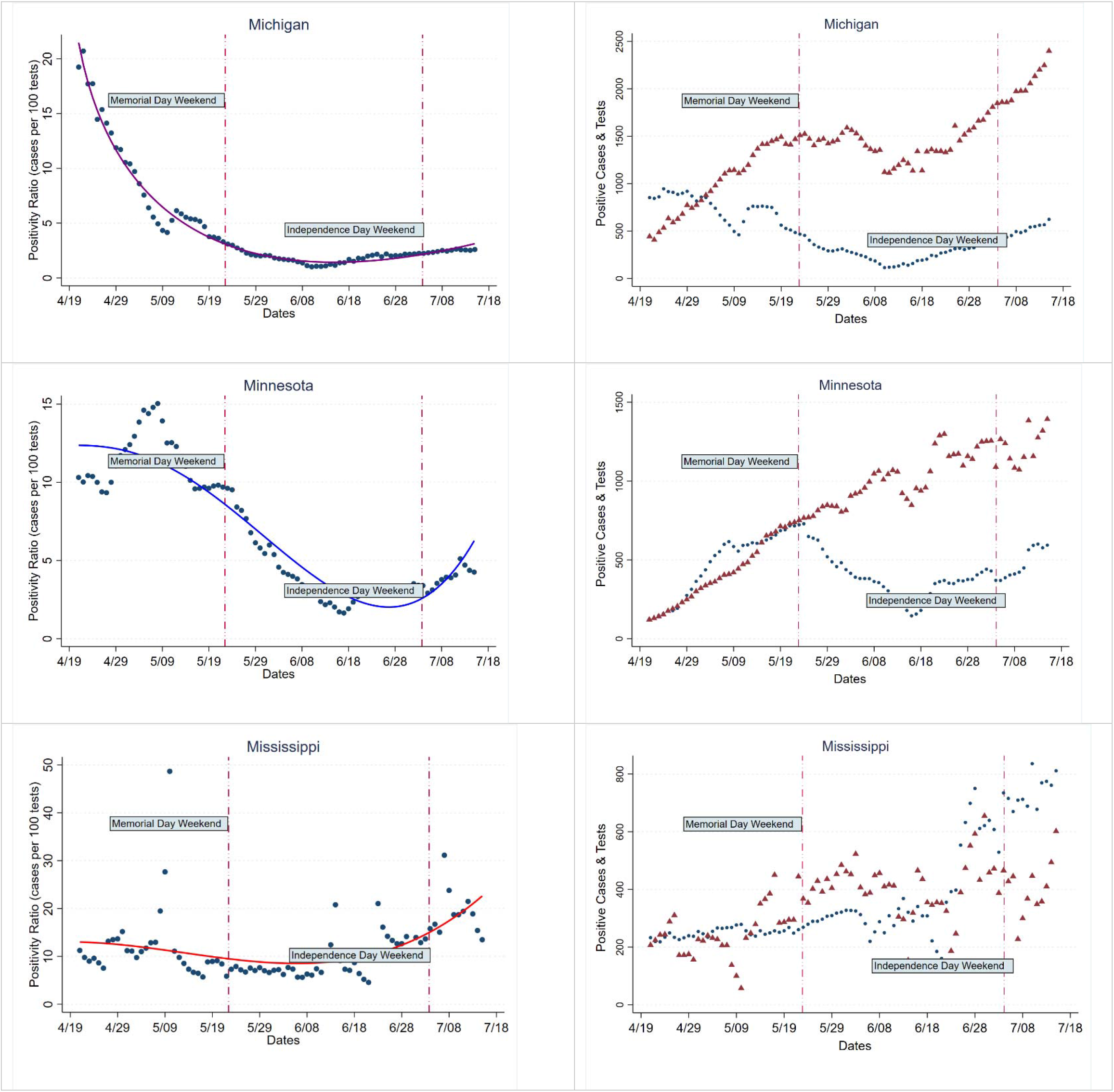
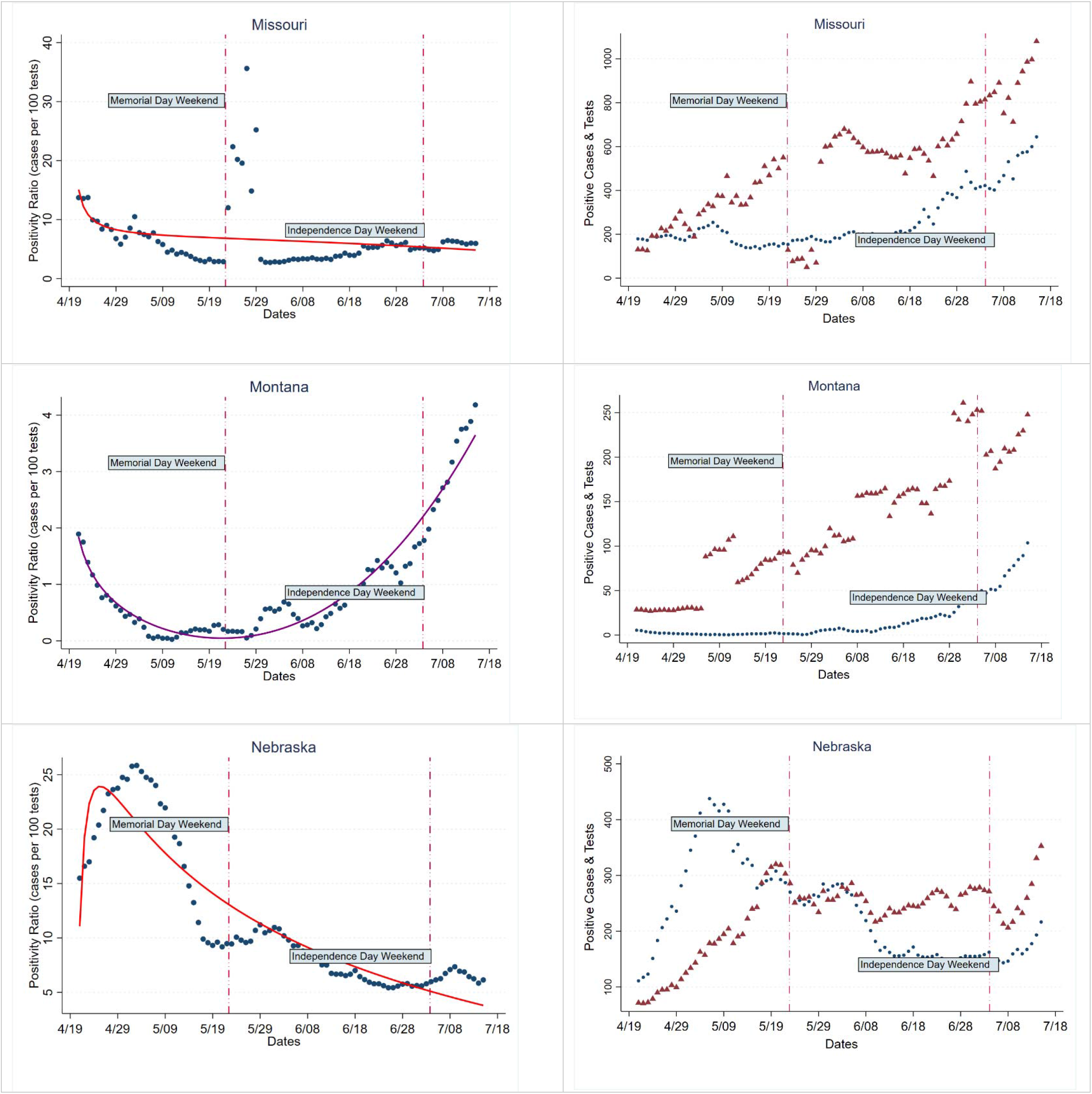
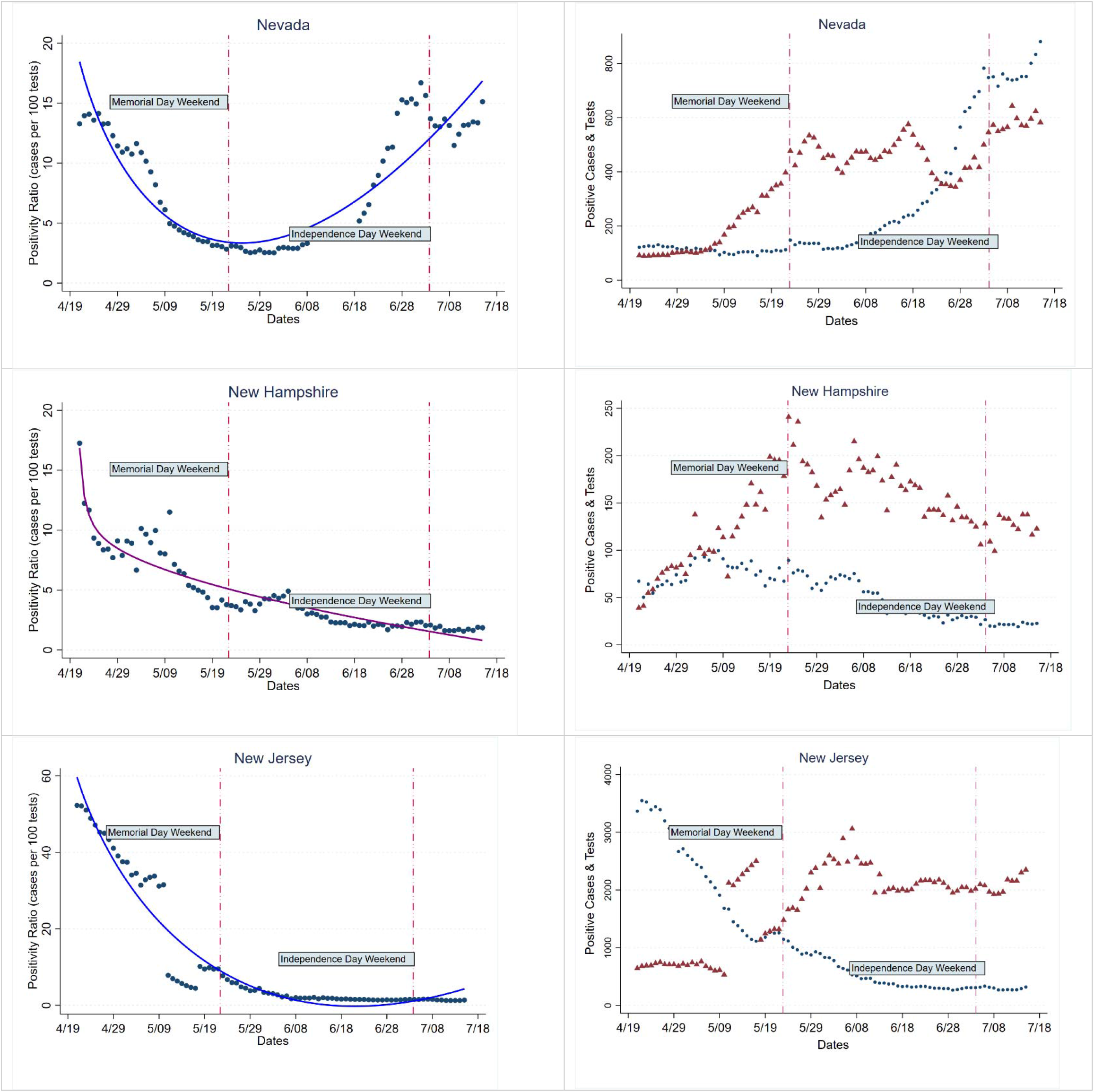
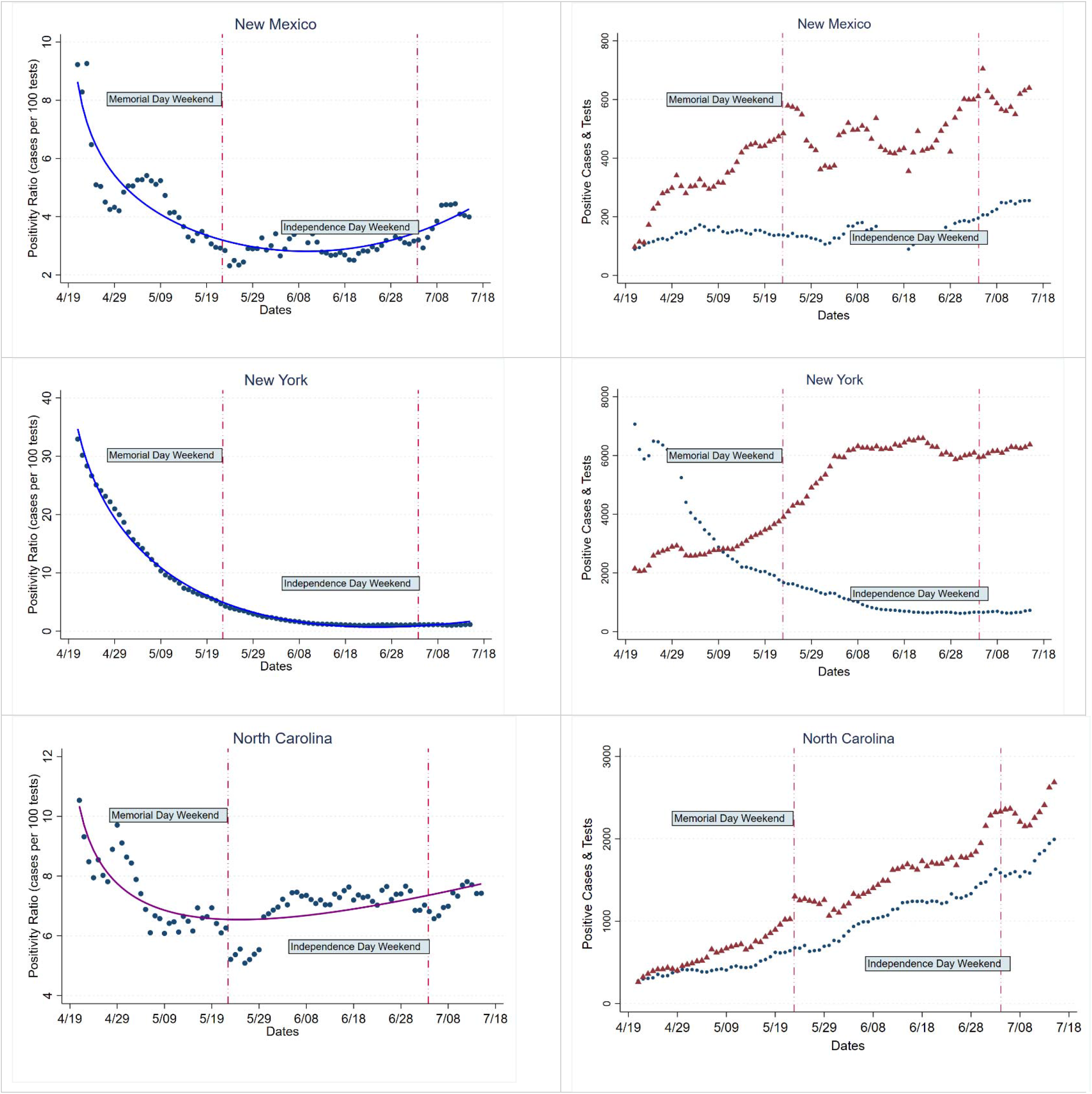
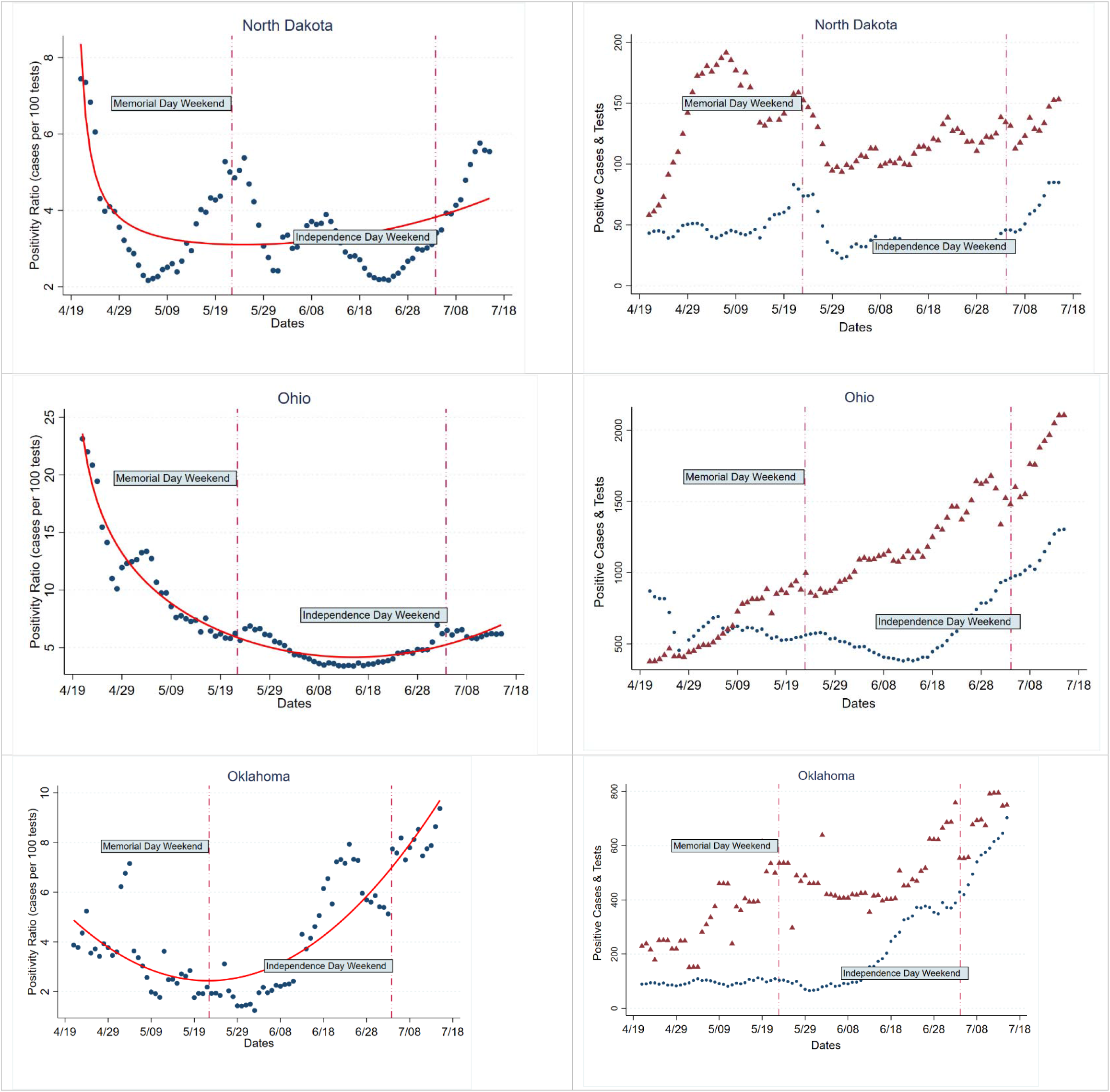
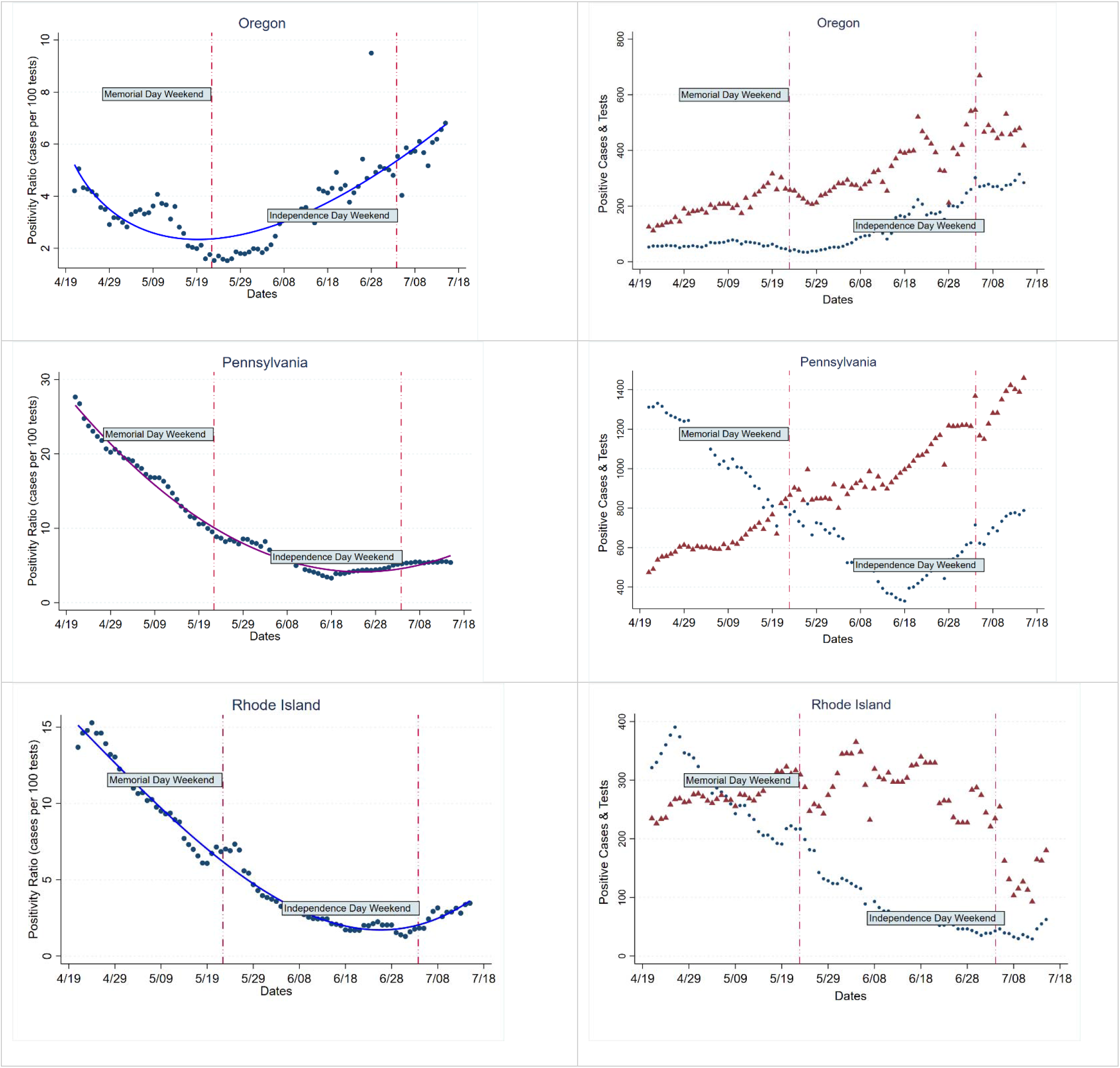
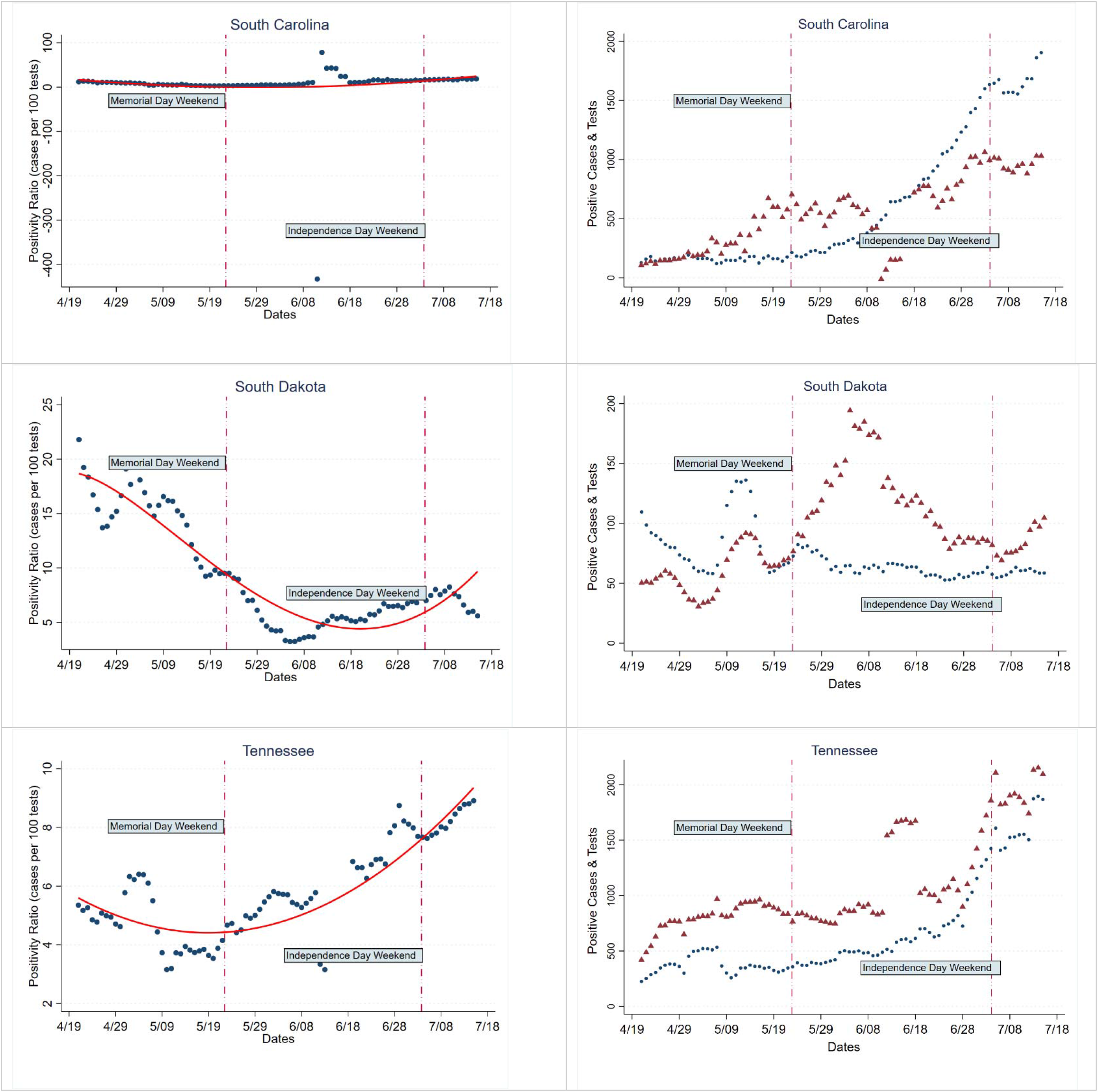
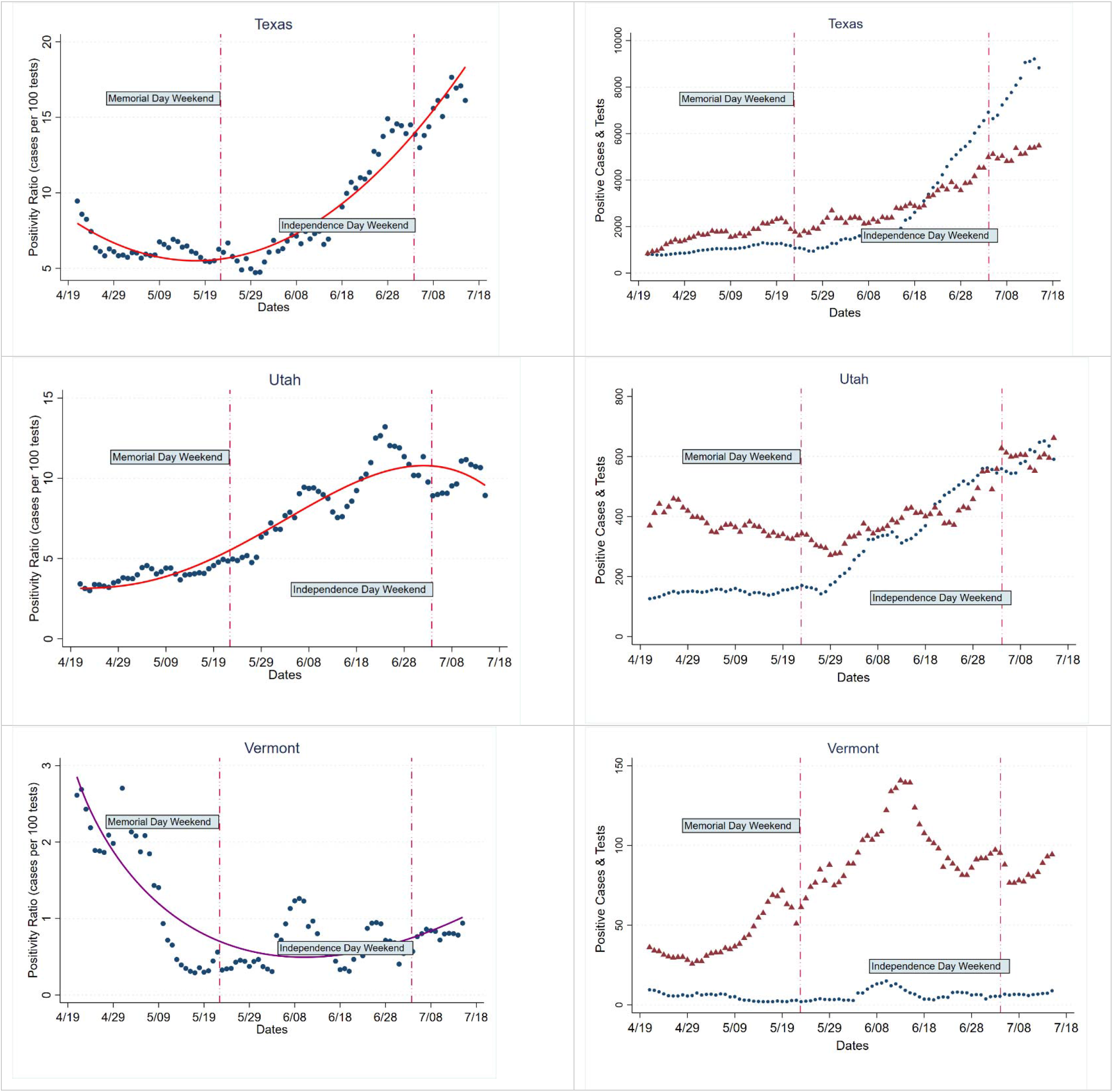
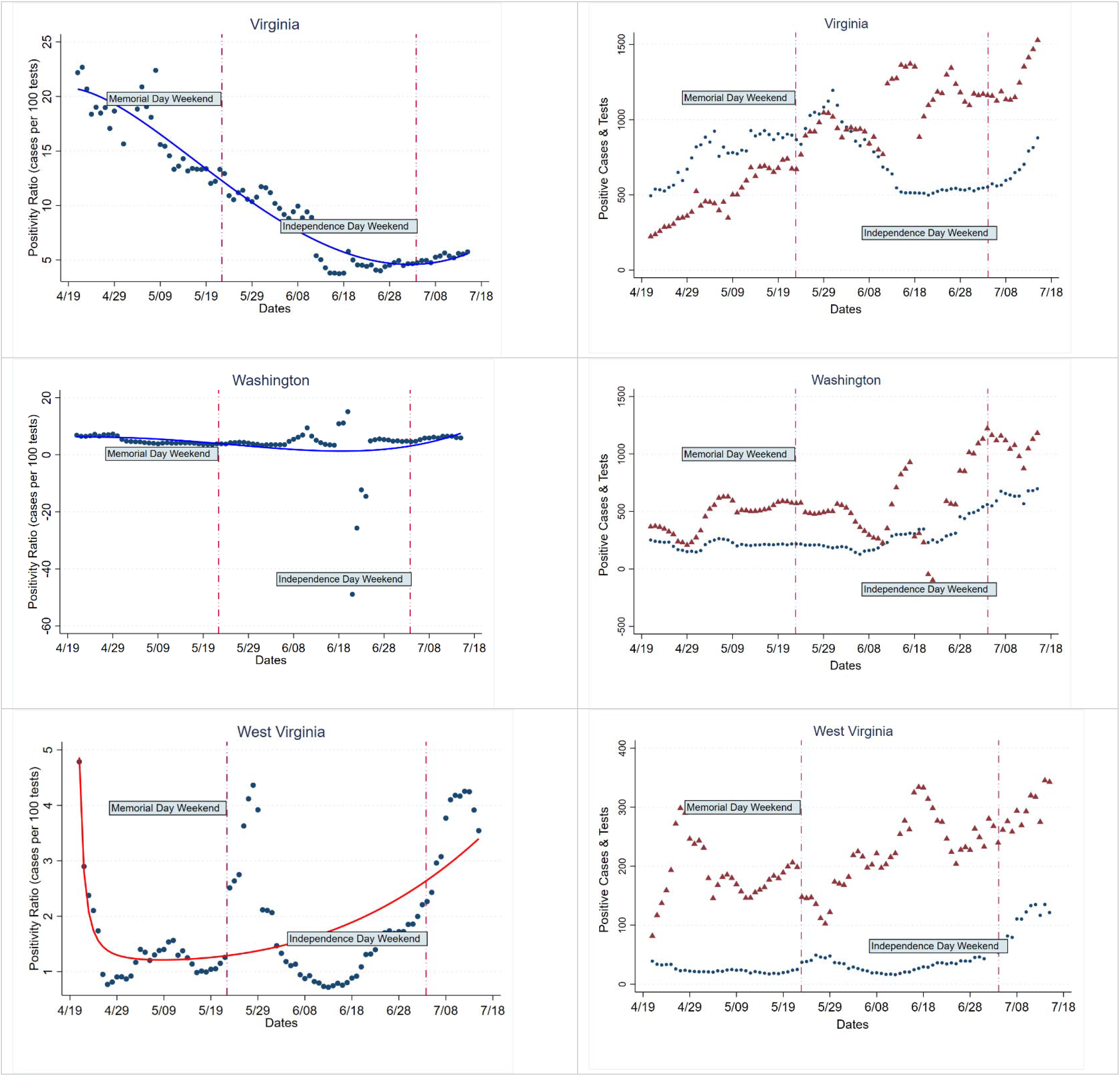
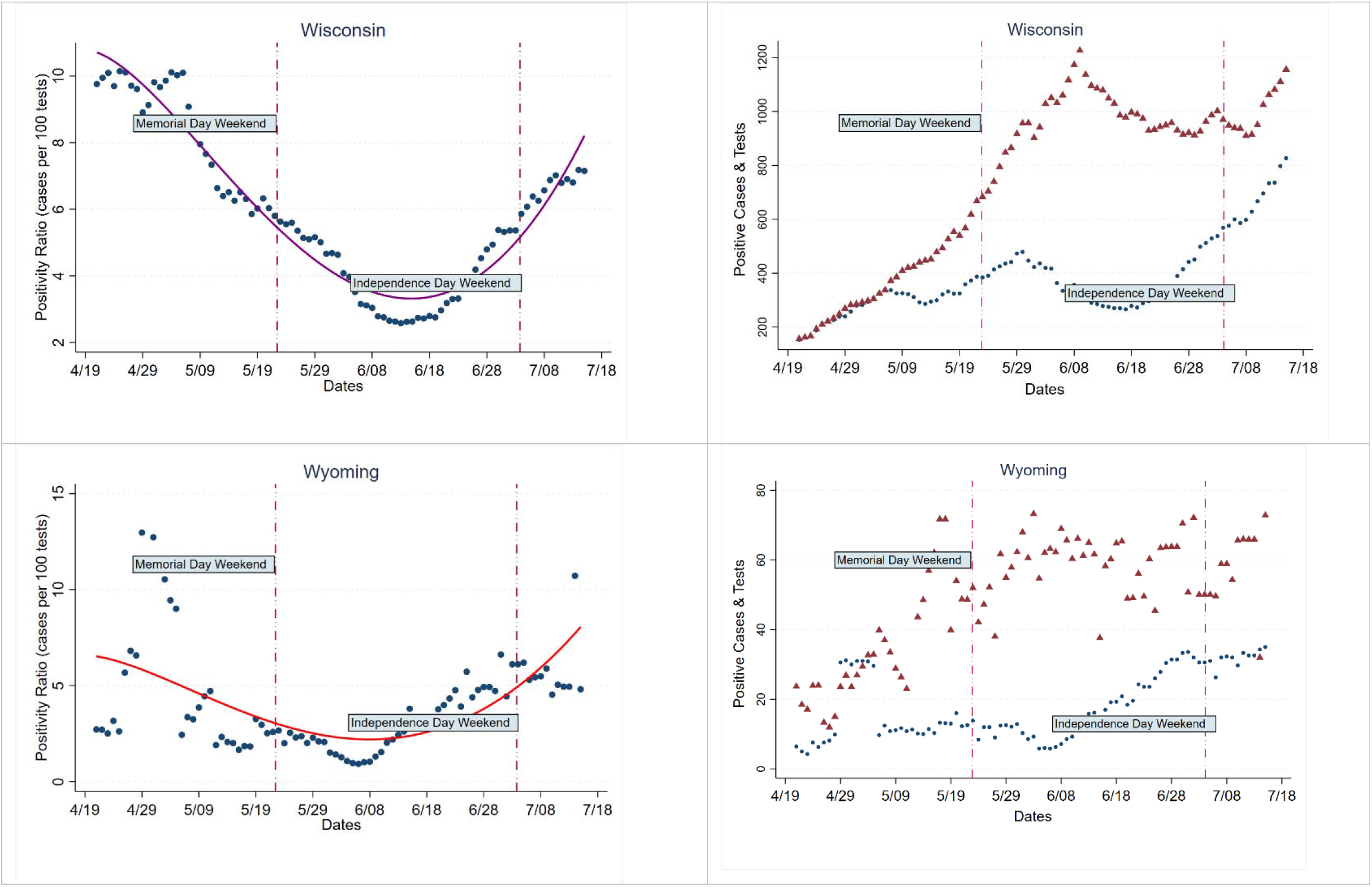
State-by-State Graphs: (1) 7-day Moving Average of Positivity Ratios with fitted trend; (2) 7-Day Moving Average of Cases & 7-Day Moving Average of Tests÷10. April 21^st^-July 15^th^, 2020. Note: ‘Negative’ numbers for tests means a correction for over-reporting on (unspecified) previous dates. We have used those negative numbers ‘as is’ instead of arbitrarily giving them a different value or dropping them.

We find that 14 states fall in Group 1, 15 states in Group 2, 15 states in Group 3, and 6 states and DC in Group 4. Republican-leaning states make up 78.57% of Group 1, 33.33% of Group 2, 40% of Group 3 and no states in Group 4 (Table 1A). The difference in political-affiliation across groups is significant (Chi2: 13.12, p<0.01). ‘High’ protest-intensity states are 64.29% of Group 1, 60.00% of Group 2, 66.67% of Group 3 and 14.29% of Group 4 (Table 1B). The difference in distribution of protest-intensity across groups is not significant (Chi2: 6.14, p>0.10). Sensitivity analyses, after excluding previously listed states with repeated negative daily test-counts, find the above results to be robust (Tables 2A and 2B).

**Table 1:** State Positivity-Ratios, Political-Affiliation & Protest-Intensity.

|  | Table 1A |  |  |  | P<br>value |
| --- | --- | --- | --- | --- | --- |
| State Type | Group1 | Group2 | Group3 | Group 4 |  |
| Republican Leaning | 11 (78.57%)<sup>a</sup> | 5 (33.33)<sup>c</sup> | 6 (40)<sup>e</sup> | 0 (0)<sup>g</sup> | .004 |
| Non Republican Leaning | 3 (21.43)<sup>b</sup> | 10 (66.67)<sup>d</sup> | 9(60.00)<sup>f</sup> | 7 (100.00)<sup>h</sup> |  |
| **Notes**<br><sup>a</sup>- MS, FL, UT, AL, TN, TX, ID, AZ, OK, SC, WY ; <sup>b</sup>- MT, OR, HI<br><sup>c</sup>- AK, GA, AR, MO, WV; <sup>d</sup>- NC, NV, LA, NM, KS, WA, WI, KY, CA, VT<br><sup>e</sup>- NE, ND, OH, SD, IN, IA; <sup>f</sup>- CO, MA, RI, MD, VA, MI, PA, MN, DE<br><sup>g</sup>- None <sup>h</sup>- NH, NJ, IL, DC, ME, NY, CT |  |  |  |  |  |
| Table 1B |  |  |  |  |  |
| BLM Protest Intensity | Group1 | Group2 | Group3 | Group 4 | P<br>Value |
| High | 9 (64.29)<sup>i</sup> | 9 (60.00)<sup>k</sup> | 10 (66.67)<sup>m</sup> | 1(14.29)<sup>o</sup> | .105 |
| Not High | 5 (35.71)<sup>j</sup> | 6 (40)<sup>l</sup> | 5 (33.33)<sup>n</sup> | 6 (85.71)<sup>p</sup> |  |
| **Notes**<br><sup>i</sup>-UT, FL, MS, TX, TN, OK, OR, AL, AZ; <sup>j</sup>- SC, WY, MT, ID, HI<br><sup>k</sup>-AR, NC, MO, GA, NV, WI, KY, CA, WA; <sup>l</sup>- NM, AK, LA, KS, VT, WV<br><sup>m</sup>-PA, VA, RI, MI, NE, OH, MD, SD, CO, MN; <sup>n</sup>- MD, IA, DE, IN, MA<br><sup>o</sup>-IL; <sup>p</sup>- DC, NJ, NY, NH, CT, ME |  |  |  |  |  |

Group1: July 13-15 positivity-ratio > Apr21-23; Group 2: Positivity-ratio in July 13-15 > Memorial Day Weekend; Group 3: Jun 29 -Jul1 < Jul 13-15. Group 4: Downward trend April 21-July15. Groups are mutually exclusive.

**Table 2:**
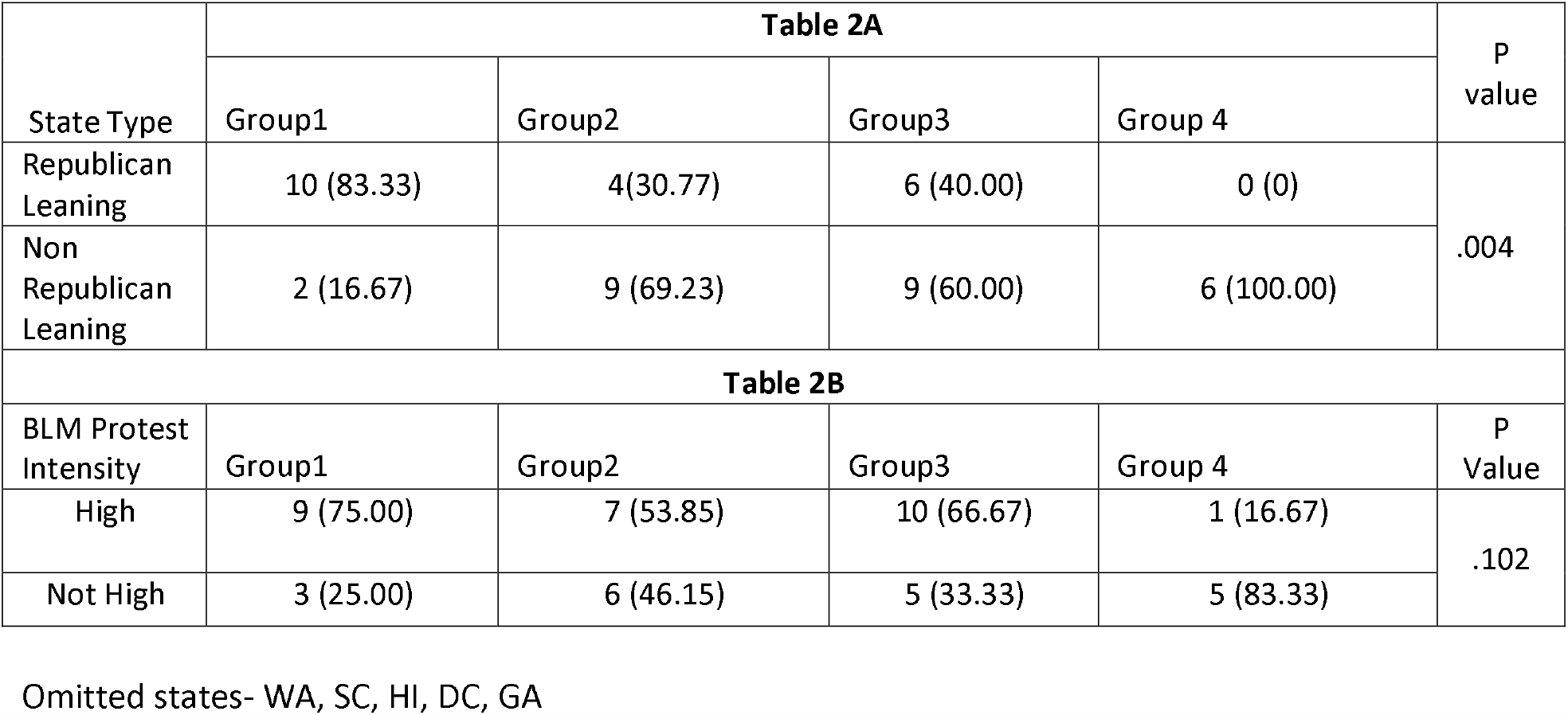
Sensitivity Analysis after Omitting States with Repeated Negative Test Counts.

## Discussion & Conclusions

Initial stay-at-home orders in U.S. states likely helped curb COVID19 infections^12,13^. It was hoped that adherence to precautionary measures would keep the disease in check after states re-opened their economies. When the initial reports of increasing cases started coming in, they were dismissed by President Trump, Vice President Pence and some other Republican leaders as an artifact of expanded testing. We believe that presenting positivity-ratios in conjunction with cases and tests allows us to discern the credibility of this argument. We present positivity-ratios from April 21-July 15 along with tests and cases for each state, and find that positivity-ratios have increased compared to late April for 14 states, compared to Memorial Day weekend for 15 states, and have started trending upwards in the last two weeks for another 15 states. This increase in positivity-ratios --even though testing has expanded beyond frontline workers and the highly symptomatic – signals a growth in underlying infection rates. Indeed, higher positivity-ratios – particularly exceeding 10%, signals that the scale of testing needs to be increased to capture the true prevalence of the disease^14^. Given this, the resistance of the current administration to support more testing is a cause for serious concern^4^.

That states with increasing positivity-ratios are disproportionately Republican-leaning is congruous with evidence of higher skepticism among Republicans about the virus^7,8^, and unwillingness to follow public-health guidance^15^. It is tempting to blame this on current Republican Party leadership; however, similar correlation of attitudes and political affiliation were found during the 2009 H1N1 outbreak^16^. There is also evidence that political affiliation is correlated with willingness to be vaccinated against diseases,^14,17^leading to the concern that, even when a COVID19 vaccine does become available, take-up may be low in Republican-leaning states. We do not find a significant association between increase in positivity-ratio and BLM protest-intensity in states, and this finding is broadly supported by more granular, city-level research that used cellphone data ^18^.

## Limitations

There are the usual concerns about quality of test and case data-reporting across states, and we find several instances of ‘negative’ daily test counts, which signals errors in earlier testing data. We further recognize that ‘political-affiliation’ can be measured in many different ways, and findings may be sensitive to this. Additionally, we recognize that our proxy measure for BLM protest-intensity may be reflective of state authorities’ attitudes towards BLM than the severity of protests per se, and more research with better methods to capture protest intensity are called for. Finally, we acknowledge that, given the fluidity of the COVID19 situation, the trends and associations that we see in this analysis may change again in the near future.

In conclusion, we find reason for serious concern about the COVID19 situation in the U.S., particularly given that we are rapidly approaching the time when decisions must be made about school-reopening. We are also concerned about the growing evidence of ‘politicization’ of the COVID19 situation, that may hamper an effective public-health response^19^. At the same time, we caution against complacency in non-Republican leaning states, given the fluidity of the disease landscape. Finally, we emphasize the need for, and strongly support increased funding for testing and adequate resources to federal-health agencies address this pandemic.

## Data Availability

All data in this manuscript are from publicly available sources.

https://covidtracking.com/

## Notes

### Competing Interest Statement

The authors have declared no competing interest.

### Funding Statement

No external funding source.

### Author Declarations

The study was approved as 'Not Human Subject' by IRB of University of Alabama at Birmingham

